# A Pilot Study on the Effects of Medically Supervised, Water-Only Fasting and Refeeding on Cardiometabolic Risk

**DOI:** 10.1101/2020.08.07.20169680

**Authors:** Eugene L Scharf, Patricia Kolbe, Su-Yeon Hwang, Natasha Thompson, Mara Gilbert, Faye Alexandrakis, Matthieu Bonjour, Evelyn Zeiler, Toshia Myers

**Author notes:** Corresponding Author:, Mayo Clinic, 200 1^st^ Street SW, Rochester, MN 55905.

## Abstract

The Fasting for Brain and Heart Health Study was a prospective, single-center, observational study examining the effects of medically supervised, water-only fasting followed by an exclusively whole-plant-food refeeding diet on accepted measures of cardiovascular risk and metabolic health. The study enrolled 48 overweight/obese, non-diabetic participants of which 26 completed the full study protocol. The participants fasted according to an established protocol at an independent medical center that reported mean fast and refeed lengths of 17 and 7 days, respectively. The primary endpoint was to describe mean glucose tolerance as indicated by Homeostatic Model Assessment of Insulin Resistance (HOMA-IR) scores at baseline, end-of-fast (EOF), and end-of-refeed (EOR) visits. Secondary endpoints were to describe the effects of fasting and refeeding on accepted markers of cardiovascular risk. Here we show that medically supervised, water-only fasting and/or whole-plant-food refeeding reduced resting systolic blood pressure (SBP), abdominal circumference, low-density lipoprotein (LDL), and high-sensitivity C-reactive protein (hsCRP) after refeeding. An increase in HOMA-IR scores at EOR was also observed.

## Introduction

Fasting is the total or partial abstention from food and/or liquid for a defined period of time that has been voluntarily practiced in religious, spiritual, and therapeutic traditions throughout history. Therapeutic fasting practices include time-restricted eating, intermittent fasting, and periodic fasting with diets ranging from low caloric to zero calorie. Practitioners of therapeutic periodic water-only fasting abstain from all caloric intake, consuming only water for between 2-40 days, in a medically supervised setting. It is worth noting that therapeutic periodic water-only fasting was found to be tolerable with a very low risk of causing a serious adverse event^1^ and that it should also be distinguished from starvation, which is an involuntary process leading to malnutrition.

Fasting causes a metabolic shift from carbohydrate metabolism to fatty acid oxidation as the primary cellular fuel source; a process termed the glucose-to-ketone (G to K) switch, which may affect numerous downstream processes^2^. Intermittent fasting has been shown to reduce circulating glucose, insulin, and insulin-like growth factor-1 (IGF-1)^3^ in animal models. Preliminary research suggests that periodic fasting results in multiple beneficial physiological effects including differential stress sensitization^4^, intracellular neuronal autophagy^5^, and upon refeeding, cellular growth, remodeling, and neuronal plasticity^2^.

Clinical research indicates that intermittent fasting may have therapeutic potential as a complementary or nonpharmacological intervention for metabolic and cardiovascular diseases^6^. A large, prospective study also recently demonstrated that periodic calorie-restricted fasting had beneficial cardiovascular and metabolic effects^7^. The effects of periodic water-only fasting followed by refeeding on cardiometabolic health are currently unknown. Here, we describe the effects of medically supervised, water-only fasting followed by refeeding on established cardiometabolic markers in participants at risk for developing cardiovascular and metabolic disease.

## Methods

### Ethical Approval

This study was approved by the Institutional Review Board at the TrueNorth Health Foundation (TNHF2018-1). The research was conducted in accordance with the approved protocol between April 2019 and February 2020 and complied with the standards of the Declaration of Helsinki. All participants provided written informed consent before data collection began.

### Participants

48 participants were recruited from patients undergoing an elective, medically supervised, water-only fast at a residential fasting center in Santa Rosa, CA, USA. Consenting participants of any sex, between 40-70 years old, with a fasting glucose <126mg/dL and/or hemoglobin A1c <7% and a BMI >25 kg/m^2^, who were approved by a non-research clinician to water-only fast for at least 10 consecutive days followed by refeed of at least 5 days were eligible for inclusion. Participant antihypertensive regimens were discontinued if admission resting systolic blood pressure was <160/100 mmHg and otherwise weaned during a whole-plant-food pre-fasting time period. Exclusion criteria included active malignancy, active inflammatory disorder including classic autoimmune connective tissue, multiple sclerosis, and inflammatory bowel disorders, and stroke or heart attack within the last 90 days.

### Medically supervised, Water-only Fasting and Refeeding Protocol

The medically supervised, water-only fasting and refeeding protocol took place a residential, integrative medical facility and was administered by non-research, clinical personnel according to previous methods^1^. The participants elected and had been approved by the attending clinician to participate in a water-only fast and refeeding before the consenting and eligibility visit. The detailed protocol is within the standards established by the International Association of Hygienic Physicians (**Supplementary Methods**). Briefly, the patients electing to water-only fast routinely underwent a comprehensive physical, neurological, and psychological examination including medical history, urinalysis, complete blood count with differentials, and comprehensive metabolic panel. Patients with contraindications (see Supplementary Methods) or taking medications that could not be discontinued, including anti-hypertensive medications and with the exception of thyroid medications which can be continued at a reduced dose, were not admitted for water-only fasting. Patients were instructed to eat a diet of fresh raw fruits and vegetables and steamed vegetables for at least 2 days before fasting. During the fast, patients remained on site, drank a minimum of 40 ounces of distilled water per day, and minimized physical activity. Medical staff monitored fasting patients twice daily and recorded vital signs and symptoms into chart notes. Urinalysis and the aforementioned blood tests were repeated weekly or as directed by a clinician. Water-only fasts were discontinued when presenting symptoms stabilized, the patient requested termination of the fast, or the clinician deemed it necessary for medical reasons. Following the fast, patients refed for a period of time lasting half of the fast length. Standard refeeding consisted of five phases (1 day per phase for each 7-10 days of water-only fasting), beginning with juice and followed by gradual introduction of solid, whole-plant foods free from added sugar, oil, and salt. Once patients were eating solid foods, the gradual reintroduction of moderate exercise was allowed. During the refeeding process, patients were offered twice-daily monitoring by clinicians.

### Study Procedures

Participants reported for study visits at baseline, every 7^th^ day of fasting and refeeding, end of fast, and end of refeed. Treatment status, resting blood pressure, weight, abdominal circumference, and 28.5mL of blood were obtained at each study visit. At baseline, demographic information and height were also obtained. Sera were sent to LabCorp for fasting glucose, insulin, hsCRP, and lipid panel analysis. The study primary endpoint, HOMA-IR, was calculated according to prior methods^19^.

### Clinical Measurements

Height (cm) was measured at the baseline visit with a wall mounted stadiometer from Doran Scales Inc. Participants were not wearing shoes. Weight, abdominal circumference, and resting blood pressure were obtained at each visit. Weight (kg) was measured using a digital body scale from Tanita (BWB 800A Class III). Participants wore a single layer of clothing but were not wearing shoes, jackets, extra sweaters, etc., and they were instructed to wear similarly weighted clothing at future visits. BMI (kg/m^2^) was calculated using the formula: weight (kg) ÷ height (metres^2^).

Abdominal circumference was measured on bare skin at the minimal waistline with a tension-sensitive, non-elastic tape (Gullick II, Model 67019). The tape was placed horizontally and parallel to the floor around the abdomen and the measurement was read on the right side of the body, in the midaxillary line, and at the end of a normal expiration.

Resting blood pressure was measured with a Connex ProBP Digital Blood Pressure Device 3400 from Welch Allyn with the participant resting and in a sitting position. Different arm cuffs were used according to the arm size: Small adult 10 for arms measuring 20-26cm (light blue cuff), adult 11 for arms measuring 25-34cm. (dark blue cuff), and large adult 12 for arms measuring 32-43cm (red cuff).

Blood was drawn in the morning, before caloric food or liquid consumption, and in a sitting position by a certified phlebotomist. Participants were instructed to drink 1-2 cups of water before the blood draw. Blood was drawn into BD Vacutainer tubes (Lavender top, 16×100, 10 ml, K2EDTA; Red top, 16×100, 10 ml, silica; Tiger top, 16×100, 8.5 ml, silica, polymer gel). The lavender top vacutainer tube was placed on ice immediately and centrifuged at 1500 g, 10 min, 4°C. The plasma was separated and frozen at -80°C. The red top and tiger top tubes were incubated at room temperature for 30 min before centrifugation at 1500 g, 10 min, 4°C. The serum was separated and was either frozen at -80°C or refrigerated at 4°C and sent to LabCorp for analysis.

### Statistical Analysis

Given the exploratory nature of the study, 25 subjects were deemed adequate to reasonably determine measures of central tendency to estimate effect sizes and plan larger clinical studies. No formal power calculations were completed. Group comparisons were performed for all subjects who completed the study protocol using two-tailed analysis of variance (ANOVA). The prespecified primary analysis was pooled unadjusted change in HOMA-IR across three paired time points: baseline, end of fast, and end of refeed assessed by repeated measure ANOVA. HOMA-IR scores were log transformed for regression modeling. Secondary analyses included changes in abdominal circumference, circulating lipid levels, and hsCRP. Exploratory analyses using unadjusted univariate linear regression were completed to determine the relationship between the fast duration and change in abdominal circumference, HOMA-IR, lipids, and hsCRP. Any finding with an alpha error of less than 5% (p<0.05) was considered significant; we did not correct for multiple comparisons. Statistical analyses were performed on JMP®, Version <14.1.0>. SAS Institute Inc., Cary, NC, 1989-2019.

## Results

### Study Population and Enrollment

Forty-eight overweight or obese, non-diabetic participants were enrolled from consenting patients at a residential medical facility specializing in water-only fasting between April 2019 and October 2019. Prior to enrollment, participants had been approved to undergo elective water-only fasting of ≥ 10 days followed by refeeding of ≥ 5 days by a non-research clinician who would supervise their entire fast. The study required that participants complete baseline (before the start of water-only fasting), EOF (at the end of at least ≥10 full days of fasting before any caloric liquids or foods were consumed), and EOR (at the end of at least ≥ 5 full days of refeed) visits to maintain enrollment. Fifty-four percent (26/48) of participants completed the full study protocol (see summary in **Figure 1**). No severe or serious adverse events were reported during fasting or refeeding. Of the 22 participants who did not complete study requirements, 9 withdrew for reasons unrelated to fasting, 1 withdrew for unknown reasons, and 12 were withdrawn because they could not complete the minimally required fast length due to adverse events. Baseline demographic and clinical characteristics of disenrolled participants as well as medical diagnoses of all participants are shown in the **Supplemental Material, Table 1**.

**Figure 1.**
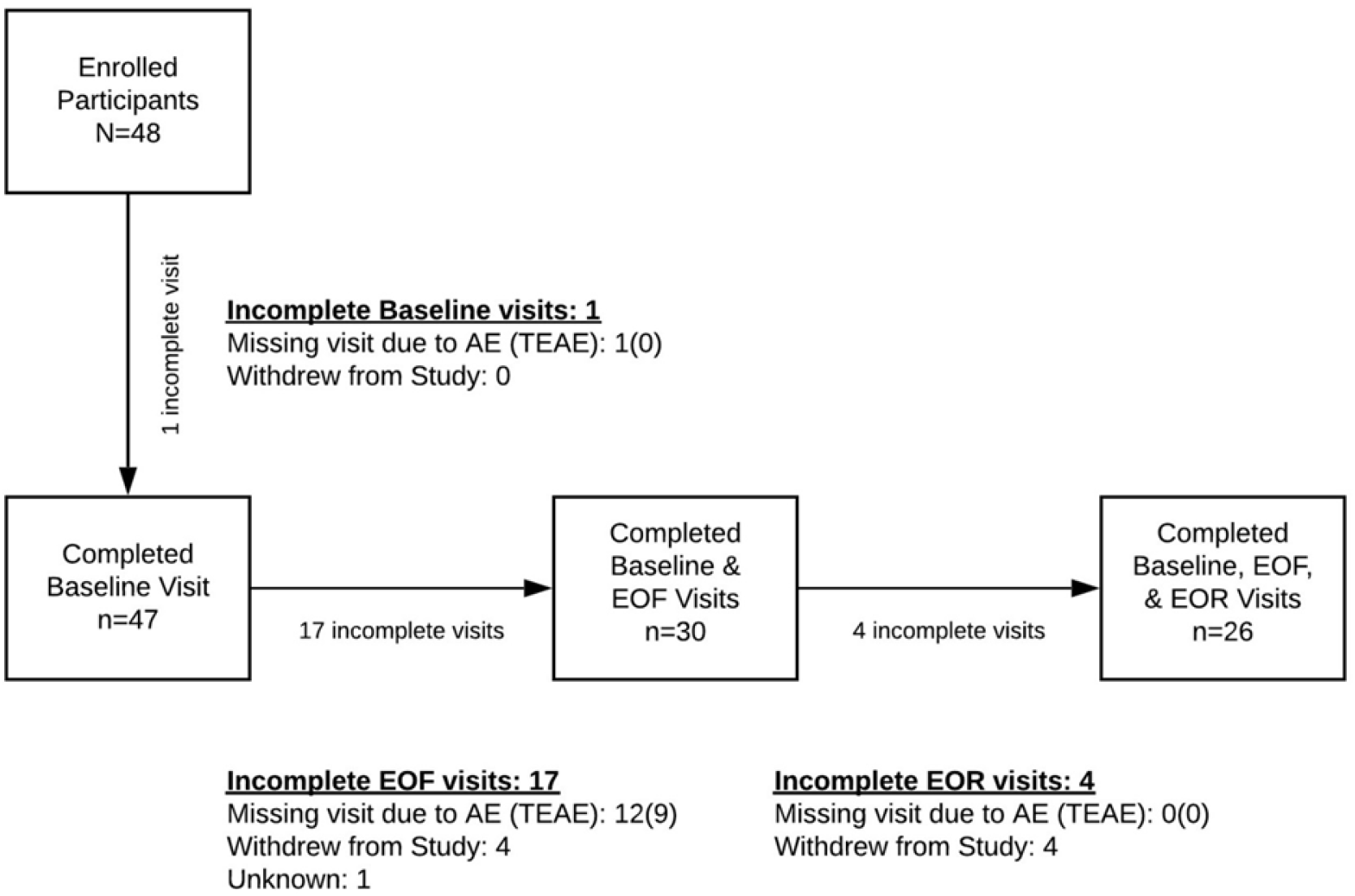
Enrollment diagram for the Fasting for Brain and Heart Health Study. EOF, end-of-fast; EOR, end-of-refeed, AE, adverse event, TEAE, treatment emergent adverse event

### Effects on Blood Pressure, Abdominal Circumference, and Body Mass Index

Baseline demographics and clinical characteristics are shown in **Table 1**. The mean fast length was 17 days and mean refeed length was 7 days. Mean systolic blood pressure decreased significantly from baseline (135mmHg) to EOF (118mmHg) and EOR (116mmHg; p=0.0002) with no change in mean resting diastolic blood pressure. Baseline abdominal circumference significantly decreased by 9cm at EOF and by 6cm at EOR (p=0.0272). A univariate linear regression showed that the reduction in abdominal circumference was associated with fast duration (r^2^=0.5, p<0.0001; **Supplemental Fig. 1**). Despite the change in abdominal circumference, there was no significant change in body mass index (BMI) during the study.

**Table 1.** The Effect of Fasting and Refeed on Cardiovascular and Metabolic Risk Factors

|  |  |  |  |  |
| --- | --- | --- | --- | --- |
| Female/Male (n) | 20/6 |  |  |  |
| Age, years, mean (SD) | 58(8)/57(10) |  |  |  |
| Mean Length of Fast, days (SD) | 17 (5.7) |  |  |  |
| Mean Length of Refeeding, days (SD) | 7 (2.6) |  |  |  |
|  | **Baseline** | **End of Fast** | **End of Refeed** | **p value\*** |
| Mean Systolic BP, mmHg (SD) | 135 (23) | 118 (10) | 116 (12) | **0.0002** |
| Mean Diastolic BP, mmHg (SD) | 81 (8.8) | 80 (7.7) | 78 (8) | 0.259 |
| Body Mass Index (BMI), kg/m<sup>2</sup> (SD) | 32 (5.4) | 29 (5.1) | 29 (5) | 0.0644 |
| Female | 32 (6) | 29 (6) | 30 (6) | 0.1546 |
| Male | 30 (3) | 27 (3) | 28 (3) | 0.2265 |
| Abdominal Circumference, cm (SD) | 99 (11) | 91 (11) | 93 (11) | **0.0272** |
| Female | 98 (12) | 89 (12) | 92 (11) | 0.064 |
| Male | 104 (8) | 97 (9) | 98 (8) | 0.355 |
| Glucose, mg/dL (SD) | 89 (10) | 76 (11) | 101 (17) | **<0.0001** |
| Insulin, μIU/mL (SD) | 8.0 (4.3) | 6.3 (4.1) | 17 (11.4) | **<0.0001** |
| HOMA-IR, | 1.8 (1) | 1.3 (0.9) | 4.5 (3.7) | **<0.0001** |
| Total Cholesterol, mg/dL (SD) | 210 (39) | 208 (50) | 188 (29) | 0.0926 |
| HDL, mg/dL (SD) | 50 (13) | 44 (9) | 44 (9) | 0.0767 |
| LDL, mg/dL (SD) | 134 (35) | 139 (50) | 108 (28) | **0.0105** |
| VLDL, mg/dL (SD) | 26 (10) | 26 (5) | 35 (9) | **<0.0001** |
| Triglycerides, mg/dL (SD) | 128 (57) | 127 (23) | 177 (43) | **<0.0001** |
| C-Reactive Protein, mg/L (SD) | 3.97 (4.4) | 4.6 (3.4) | 2.38 (1.9) | 0.0611 |

### Effects on glucose, insulin, and HOMA-IR

The primary endpoint was to assess mean HOMA-IR scores [fasting insulin (microU/L) x fasting glucose (nmol/L)/22.5] after fasting and refeeding. Consistent with prior literature^8-10^, fasting alone significantly reduced blood glucose and insulin values compared to baseline, but both values were significantly increased at EOR compared to baseline (**Table 1**). Accordingly, the mean HOMA-IR score was significantly elevated at EOR (4.5; p<0.0001) compared to baseline (1.8) and EOF (1.3) (**Table 1**). Baseline HOMA-IR scores were strongly correlated with EOR HOMA-IR scores (r^2^ =0.55, p<0.001; **Figure 2**), although the increase from baseline to EOR was not associated with the duration of fast or refeed (data not shown; p=0.0612 and p=0.13, respectively). **Figure 3** shows an overview of individual HOMA-IR scores at baseline, EOF, and EOR separated by fast duration.

**Figure 2.**
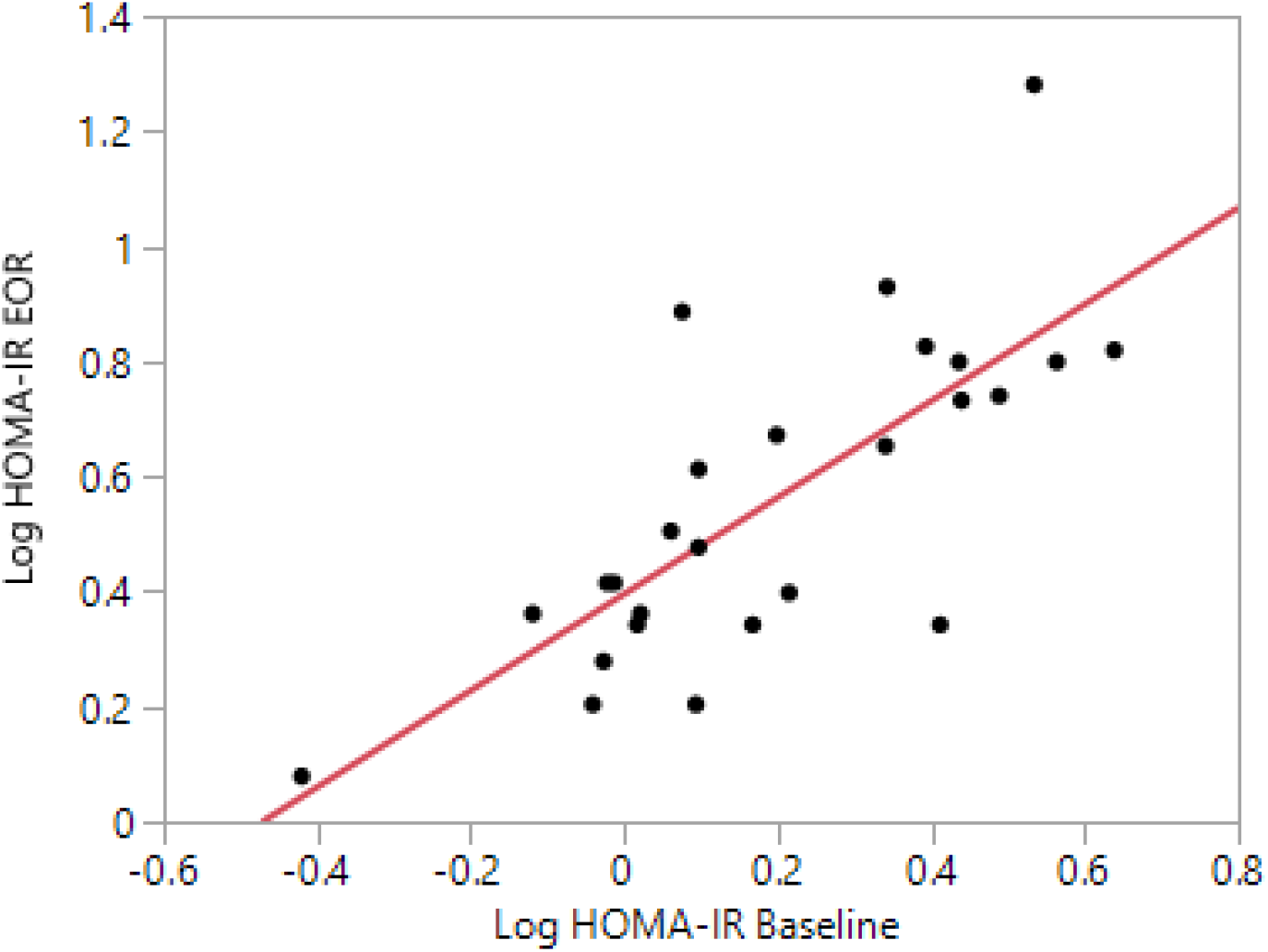
HOMA-IR Baseline and End of Refeed. End of refeed HOMA-IR was significantly associated with baseline measure. HOMA-IR values log transformed.

**Figure 3.**
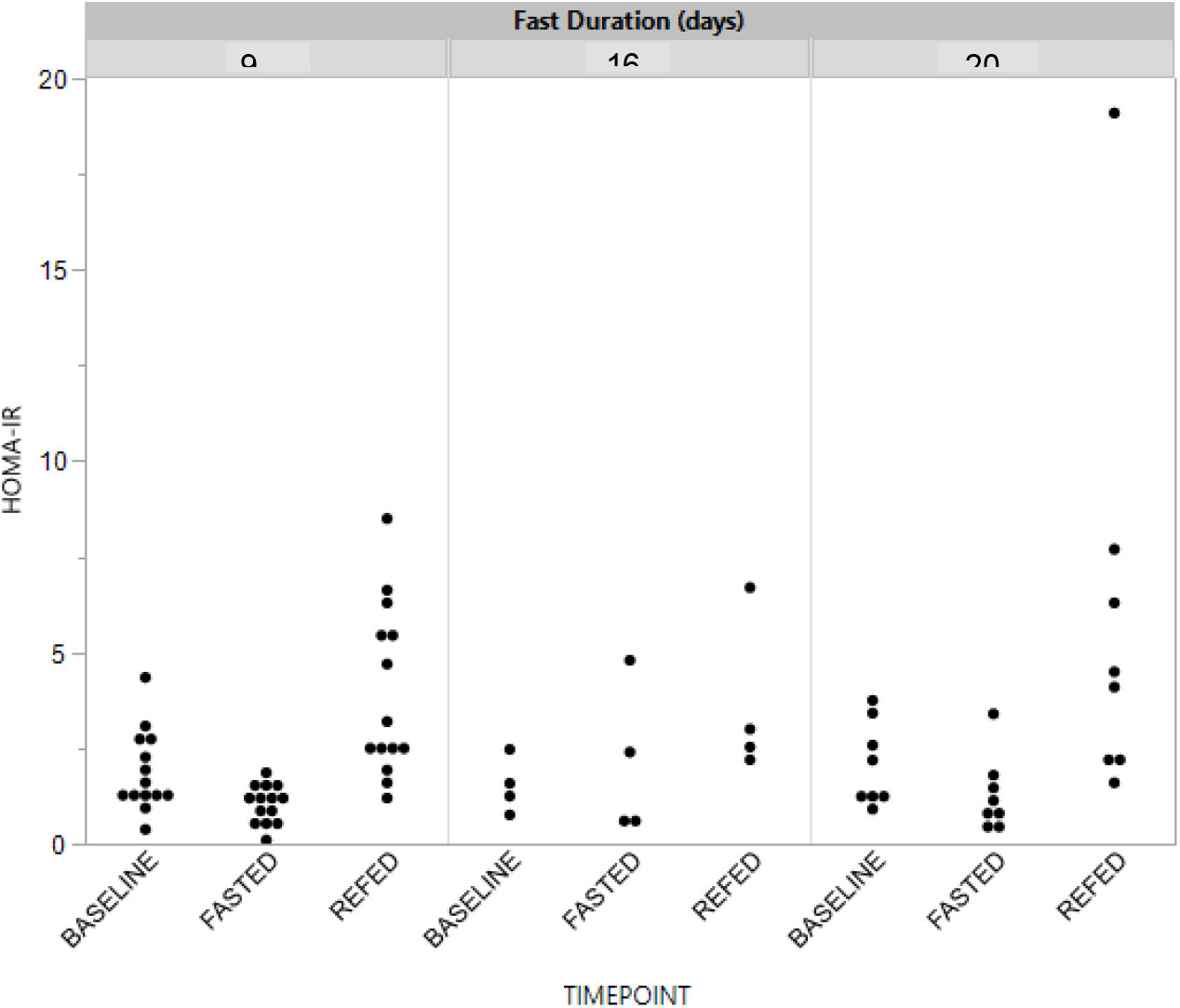
Homeostatic Model Assessment of Insulin Resistance (HOMA-IR) During Medically Supervised, Water-Only Fasting and Refeeding. HOMA-IR scores are shown at baseline, end-of-fast (fasted), and end-of-refeed (refed) visits across different fast duration tertiles.

### Effects on Serum Lipoproteins

The effects of water-only fasting and refeeding on serum low-density lipoprotein (LDL) are shown in **Figure 4**. Paired mean change in LDL between baseline and EOR time points was - 26mg/dL (133mg/dL v 107mg/dL, p<0.0001; **Figure 4A**). The mean change for participants with baseline LDL >120mg/dL (n=16) was -34mg/dL (156mg/dL v 122mg/dL, p<0.0001; **Figure 4B**). Notably, fast duration did not have an effect on the change in LDL (**Figure 4C**). Mean change in total cholesterol (**Table 1**) was correlated with the change in LDL (r^2^=0.74, p<0.0001; **Figure 4D**). There was no significant change in high-density lipoprotein (HDL; **Table 1**). Mean very-low-density lipoprotein (VLDL) and triglyceride levels increased significantly from baseline to end-of-refeed time points from 26 mg/dL to 35mg/dL (p<0.0001) and 128mg/dL to177mg/dL (p<0.0001), respectively (**Table 1**).

**Figure 4.**
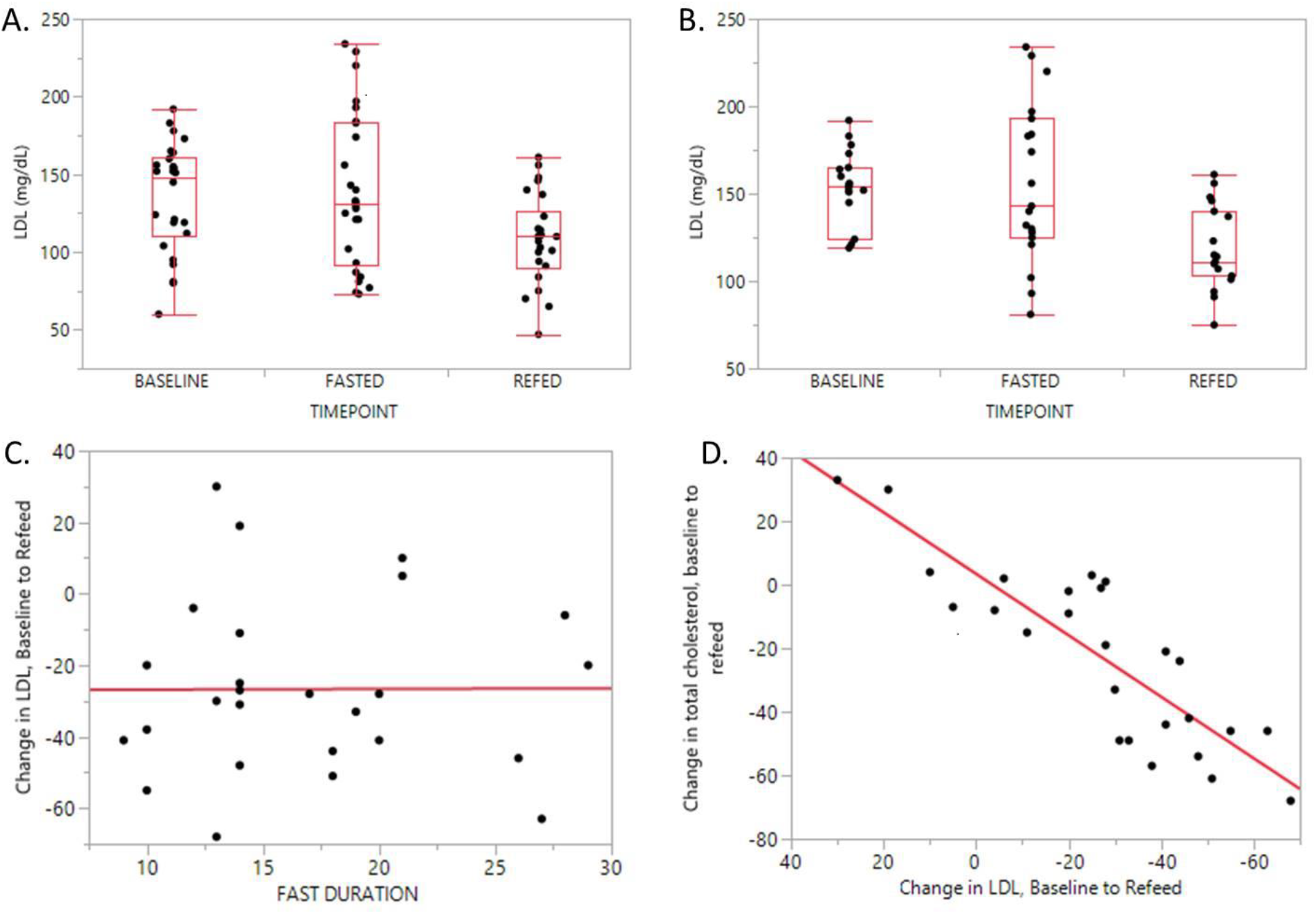
Serum Low-Density Lipoprotein (LDL) Response to Water-Only Fasting and Refeeding. **A**. LDL (mg/dL) at baseline versus end-of-refeed, mean 135mg/dL v 108mg/dL, p=0.0021 **B**. LDL (mg/dL) at baseline versus end-of-refeed for participants with baseline LDL>120. 156mg/dL v 123mg/dL, p<0.001 **C**. Change in LDL as a function of Fast Duration, p = 0.98, r2 =1.83×10^-5^ **D**. Change in Total Cholesterol as a function of Change in LDL, p <0.0001, r^2^= 0.74

### Effects on high-sensitivity CRP

The mean change in serum hsCRP from baseline to EOR was -1.6mg/L (4.0 mg/L v 2.4 mg/L, p=0.06; **Table 1**). In participants with baseline hsCRP levels >2mg/L, baseline to EOR paired mean change was -4.6mg/L (6.02mg/L v 1.44mg/L, p = 0.013, n=17). Mean serum hsCRP levels increased from baseline to the EOF, but this finding was not significant (**Table 1**). Fast duration was not associated with the decrease in hsCRP levels. However, the refeed duration was associated with the decrease in hsCRP levels at the EOR time point (r^2^= 0.21, p=0.0173; **Supplemental Fig. 2**).

## Discussion

This study describes the effects of water-only fasting followed by an exclusively whole-plant-food refeed diet on insulin resistance, serum lipoproteins, and inflammation. Our results showed a significant increase in HOMA-IR scores after refeeding that were not associated with any fast length but were positively associated with HOMA-IR scores at baseline. These findings suggest that any fast of 10 days or longer may result in insulin resistance upon refeed in overweight and obese individuals and that baseline glucose intolerance increases the risk of insulin resistance during the refeeding period immediately after fasting ends.

Increased HOMA-IR scores upon refeeding may be a rebound phenomenon as the metabolic switch reverses from ketones back to glucose as the primary fuel source. Insulin receptor expression has been shown to downregulate during longer periods of fasting^11^, but the effects of refeeding on insulin signaling have not yet been studied. Our data show that insulin levels increase during refeeding, which may signify insulin resistance. This finding is supported by the elevated triglycerides and VLDL after refeeding, which may further interfere with insulin signaling and lead to a transient “pseudo-insulin-resistant state” during the metabolic switch from ketosis to glycolysis^9^. Longer-term follow-up studies are underway to confirm our findings and determine whether increased insulin resistance is a transient metabolic phenomenon, as previously suggested^9^. Recent literature demonstrating favorable cardiometabolic effects of periodic fasting have not reported post-refeed fasting glucose and insulin levels^12^. Therefore, it would also be interesting to assess glucose and insulin levels during the refeeding period of other fasting methods. Expanding our analyses to include a broader range of fasting lengths as well as metabolically normal participants may provide additional insights into the physiology of insulin resistance as well as the potential health benefits of this intervention.

This study is consistent with previous data suggesting that water-only fasting has a rapid antihypertensive effect that persists after refeeding^13^. Decreased blood pressure may be the result of natriuresis observed during fasting, decreased sympathetic activation, and/or favorable changes in the sensitivity of the renin-angiotensin-aldosterone system (RAAS). No cases of electrolyte disturbance were reported and patients were hydrated *ad libitum* during the study making hypovolemia a possible but less likely explanation. An ongoing study will examine the neurohumoral effects of water-only fasting and whole-plant-food refeeding in order to gain a greater understanding of the antihypertensive effects of this intervention and whether the fast has long-term residual effects on RAAS sensitivity.

Serum LDL levels were significantly decreased after refeeding compared to baseline particularly in participants with elevated baseline measures. This may be due to increased LDL receptor activity, likely from cholesterol-deficient hepatocytes resulting from the fast. However, LDL was not significantly changed at EOF, suggesting that hepatocyte clearance of circulating cholesterol did not occur until feeding resumed. The phytosterol content of the whole-plant-food diet consumed during refeed may have contributed to the observed reduction in LDL values by stimulating hepatic LDL receptor mediated clearance. Although the degree of LDL reduction observed in this study is modest compared to pharmacotherapy for hypercholesterolemia, the effect is notable for a one-time intervention and suggests the need for long-term follow-up studies to determine the duration of this effect, whether repeated cycles may have additive effects, and whether the effect is explained by the dietary intervention alone.

CRP is an acute-phase-reactant marker of inflammation and metabolic stress^14^. Prior reports describe elevated CRP during calorie-restricted fasting^12,15^, which was attributed to circulating catecholamines. We did not observe a significant change in hsCRP after fasting. However, after refeeding, hsCRP was significantly lower in participants with elevated hsCRP at baseline. Notably, refeed duration was positively correlated with decreased hsCRP levels prompting the question as to whether the whole-plant-food diet is responsible for the anti-inflammatory effects observed in this study. These findings suggest a hormetic effect on inflammation and require further investigation.

This study recruited from patients who were voluntarily undergoing medically supervised, water-only fasting at a residential medical center. The study protocol required participants to complete at least 10 full days of fasting, at least 5 full days of refeeding, all study visits, and the EOF blood draw before breaking the fast. Ultimately, 46% (22/48) of participants were disenrolled because they did not meet one or more of these requirements. The low retention rate raises issues about the tolerability of water-only fasting. However, only 25% (12/48) of participants were unable to complete the study requirements because their fast was prematurely broken or suspended due to adverse events possibly attributable to fasting, all of which were mild to moderate and known to occur with fasting as previously reported^1^. The low retention rate is more likely a reflection of the practical difficulties of conducting water-only fasting research in humans, such as on-site participant time investment (e.g., it may not be feasible for subjects to spend three weeks completing the intervention). Furthermore, in clinical practice, early termination or temporary suspension of a fast with juice intake is not necessarily an indicator that the fast was intolerable or unsuccessful. Nonetheless, water-only fasting has been criticized for difficulty with patient tolerance^16^, which has resulted in various modified fasting protocols that allow a minimal amount of caloric intake <500kcal per day while preserving ketosis (e.g., fasting mimicking diet^17^ and Büchinger fast^18^). Whether a scientifically meaningful difference in health benefits exists between these protocols (water-only vs. calorie-restricted fasting) has not been rigorously investigated and should be pursued in randomized trials.

The optimal duration of fasting is currently unknown. The study protocol described the effects of fasts ≥10 days because the length is considered minimally sufficient for the population based on clinical practice from thousands of patients. Nonetheless, there have not been any clinical studies to determine the optimal duration of a periodic water-only fast, which likely varies based on baseline characteristics of the subject and purpose of the therapeutic fast. For example, the observed reduction in LDL was not correlated with fast duration while the reduction in abdominal circumference was strongly correlated. In the future, it will be necessary to have larger samples sizes and a broader range of fast lengths (i.e., 2-40 days) to address this question.

A major strength of this study is that the water-only fasting and refeeding protocol was supervised in a domiciled setting, which should have minimized fasting and dietary noncompliance and permitted the assessment of participants who water-only fasted 10 or more days. Another strength of this study is that it is the first to report on the metabolic effects that occur during refeeding. This study also has several limitations including that it is a single-center study with a small sample size and therefore external validity cannot be assessed. There is also a referral bias in that we recruited participants who were already intending to fast, which could potentially reflect more motivated patients and limits external validity. An additional limitation is the lack of long-term follow-up visits, which is currently being addressed in an ongoing study.

## Conclusion

The Fasting for Brain and Hearth Health Study suggests that periodic water-only fasting and exclusively whole-plant-food refeeding may be an effective, nonpharmacological intervention for patients with cardiometabolic risk factors such as increased abdominal circumference, elevated LDL cholesterol, or elevated CRP. However, refeeding after periodic water-only fasting may temporarily induce an insulin resistant state. Studies are currently underway to determine if this is a transient rebound phenomenon.

## Data Availability

The data that support the findings of this study will be available upon request after peer-reviewed publication.

## Acknowledgements

The authors would like to thank Alan Goldhamer and the medical staff at TrueNorth Health for providing the clinical site at which this research was conducted.

**Abbreviations**
CRP: C-reactive protein
HDL: high-density lipoprotein
HOMA-IR: Homeostatic model of insulin resistance
LDL: low-density lipoprotein
VLDL: very-low-density lipoprotein
EOF: end-of-fast
EOR: end-of-refeed

## Notes

### Competing Interest Statement

The authors have declared no competing interest.

### Clinical Trial

Not an applicable clinical trial.

### Funding Statement

No external funding was received.

### Author Declarations

This study was approved by the Institutional Review Board at the TrueNorth Health Foundation (TNHF2018-1).

